# Developing a Unified Criminal Justice Pathway into Drug and Alcohol Treatment from Police Custody: A Public Health Service Evaluation and Pathway-Design Project in Blackpool, United Kingdom

**DOI:** 10.64898/2026.06.07.26355095

**Authors:** Abubakar Badmos, Ahmed O. Abdulkareem, Ann Gawne, Judith Mill, Toyeeb Idris

## Abstract

**Introduction:** Blackpool, England’s most deprived local authority, has the highest drug-related death rate in the country. People in police custody with problem substance use are a key Core20PLUS5 inclusion-health group, yet referral from the police into structured drug and alcohol treatment is fragmented and relies heavily on self-report. We evaluated the current police-to-treatment route in Blackpool and designed an evidence-informed unified pathway.

**Materials and Methods:** A mixed-methods service evaluation and pathway-design project was conducted during a six-month General Practice / Public Health rotation. Routinely collected referral data from Horizon (the local specialist drug and alcohol service) covering the 47-month period December 2019 – October 2023 were analysed. Findings were triangulated with national policy, the Project ADDER and Liaison and Diversion evaluations, and the international evidence on police-led pre-arrest diversion.

**Results:** Of 5,900 total referrals into Horizon over 47 months, only 269 (4.56%) originated from the police. Police referrals accounted for fewer than 5% of monthly referrals in 30 of 47 months, for 5-9.9% in 16 months, and for ≥10% in only one month (10.8%, December 2022). Blackpool recorded 76 drug-misuse deaths in 2019 is 21 (19.4 per 100,000, approximately four times the England rate). A six-step unified pathway is proposed: Initiate Referral (opt-out, from ADDER Police and Liaison and Diversion); Initial Assessment; Tailored Treatment Plan; Continuous Support; Collaboration and Monitoring; and Evaluation and Adjustment.

**Conclusions:** Police contact is markedly under-used as a gateway to treatment despite Blackpool having the highest drug-related mortality in England. An opt-out, multi-agency pathway anchored in Core20PLUS5 has the potential to narrow the treatment gap, reduce re-offending, and address the structural health inequalities that drive premature mortality.

**Citation:** Abdulkareem AO, Badmos A, Gawne A, Mill J, Idris T. Developing a unified criminal justice pathway into drug and alcohol treatment from police custody: a public health service evaluation and pathway-design project in Blackpool, United Kingdom.

## 1. Introduction

Substance misuse is one of the strongest drivers of premature mortality, morbidity and criminal-justice involvement in the United Kingdom. In 2022, there were 4,907 drug-poisoning deaths registered in England and Wales, the highest number since comparable records began in 1993, and the rate has continued to rise in subsequent years [1,2]. Drug-related deaths cluster sharply in the most deprived communities and in coastal towns, where economic decline, poor-quality transient housing and the contraction of public services interact to exacerbate vulnerability [3,4].

Blackpool exemplifies this pattern. The town has been ranked the most deprived local authority in England on both rank-of-average-rank and rank-of-average-score measures since the 2019 English Indices of Deprivation, a position retained in the 2025 update [4,5]. Approximately 40% of Blackpool’s Lower-layer Super Output Areas (LSOAs) fall within the 10% most deprived nationally, and several of the ten most deprived neighbourhoods in England are within its boundaries [4]. Male life expectancy in Blackpool is 5.8 years below the England average, and male healthy life expectancy is the lowest of any local authority in England [6,7]. Drug and alcohol misuse is a principal contributor to this gap, alongside smoking and cardiovascular disease.

Against this backdrop, Blackpool has had the highest drug-related death rate in England for much of the last decade, with an age-standardised mortality rate of 19.4 per 100,000 in 2019–21 (76 deaths), approximately four times the contemporaneous England rate of approximately 5.1 per 100,000, and more recent figures showing further deterioration [1,8]. Blackpool was designated one of the five original pilot sites for Project ADDER (Addiction, Diversion, Disruption, Enforcement and Recovery), the Home Office-led, whole-system programme launched in 2020 and extended as part of the 2021 Drug Strategy, *From Harm to Hope: A 10-year Drugs Plan to Cut Crime and Save Lives* [9,10]. ADDER was explicitly designed to combine targeted enforcement with enhanced diversion, treatment and recovery provision in the areas of England and Wales worst affected by drug-related harm [10,11].

Within the NHS, the Core20PLUS5 framework published by NHS England in 2021 provides a structured approach to reducing healthcare inequalities. It directs accelerated improvement at the most deprived 20% of the population (Core20), locally determined inclusion-health groups (PLUS), which explicitly include people experiencing homelessness, drug and alcohol dependence and those in contact with the justice system, and five priority clinical areas [12,13].People in police custody with problematic substance use sit at the intersection of multiple Core20PLUS groups and are therefore a priority target population for healthcare-inequality interventions.

Despite this policy convergence, referral from the police into structured treatment has historically been the weakest limb of the English criminal-justice-to-treatment pathway. Evaluations of arrest-referral schemes from the early 2000s, of Liaison and Diversion (L&D) services from 2014 onwards, and of Project ADDER itself have all identified variable implementation, inconsistent uptake and persistent reliance on self-report as structural limitations [11,14,15]. In Blackpool, the principal established referral routes into the specialist drug and alcohol service (Horizon), involving prison release, probation, court, the Lived Experience Team and self-referral, function reasonably well, but referral from L&D and the ADDER Police team is widely perceived by local stakeholders to be fragmented, producing low treatment uptake and high re-offending among a population at exceptionally high risk of premature death.

This project was commissioned by the Blackpool Public Health Directorate and undertaken as part of a six-month General Practice / Public Health rotation. Its objectives were:

- to quantify the current contribution of the police to treatment referrals in Blackpool;
- to identify the structural and operational barriers to police referral;
- to propose a workable, evidence-informed unified pathway that could be embedded in routine multi-agency practice; and
- to situate this pathway within the Core20PLUS5 health-inequalities framework and the legacy of Project ADDER.

## 2. Materials and Methods

### 2.1 Design

This was a mixed-methods service evaluation and pathway-design project. It combined a descriptive analysis of routinely collected referral data with a narrative review of relevant policy, service evaluation, and peer-reviewed literature.

### 2.2 Setting

Blackpool is a coastal unitary authority in North-West England with a resident population of approximately 139,000. Specialist adult drug and alcohol treatment is commissioned from Horizon, with complementary services provided by the Adelaide Street Service, the OASIS partnership, the Women’s Centre, the Lived Experience Team, the Lighthouse Project, and community counselling providers. Police custody in Blackpool is served by Lancashire Constabulary, with criminal-justice healthcare input through the Liaison and Diversion team and, during the study period, the Project ADDER Police team.

### 2.3 Data sources

Quantitative data were extracted from Horizon’s routine referral monitoring returns for the 47-month period December 2019 – October 2023, covering all recorded new treatment referrals by source (prison, probation/court, Lived Experience Team, self-referral, police/L&D, other). Monthly totals and the number and proportion originating from the police were tabulated.

Contextual epidemiological data on deprivation, life expectancy, drug-related deaths and drug-related hospital admissions were drawn from the Blackpool Joint Strategic Needs Assessment (JSNA), the Office for National Statistics (ONS) drug misuse mortality release, the Office for Health Improvement and Disparities (OHID) Fingertips tool, and the Ministry of Housing, Communities and Local Government’s English Indices of Deprivation 2019.

### 2.4 Pathway development

Pathway development followed an iterative approach. Findings from the referral-data review were triangulated with (a) the published Project ADDER evaluations [11,16], (b) the RAND Europe evaluation of the national Liaison and Diversion programme [14], (c) the international literature on opt-out and police-led pre-arrest diversion to treatment [17,18], and (d) stakeholder discussions with the Harm Reduction Lead, the Public Health Consultant, and clinical and operational contacts within Horizon, Lancashire Constabulary ADDER and the Liaison and Diversion service. Pathway components were mapped against the Core20PLUS5 framework to ensure explicit alignment with NHS priorities for healthcare-inequality improvement [12,13].

### 2.5 Statistical analysis

Monthly referral counts were summarised descriptively. The number of months in which police referrals accounted for <5%, 5–9.9%, and ≥10% of total referrals was tabulated, and the maximum monthly police-referral share was identified. No inferential statistical testing was performed; the purpose of the quantitative analysis was to characterise magnitude and pattern rather than to test hypotheses. All analyses were conducted in Microsoft Excel (Microsoft Corporation, Redmond, WA, USA).

### 2.6 Ethical considerations

This project was a public-health service evaluation commissioned by the Blackpool Public Health Directorate and conducted under the Directorate’s routine clinical-governance and service-improvement arrangements. The activity was classified as a service evaluation rather than as research. This determination was made by the supervising Public Health team at Blackpool Council, specifically Ms Ann Gawne (Harm Reduction Lead) and Dr Judith Mill (Consultant in Public Health), applying the criteria set out in the UK Health Research Authority (HRA) decision tool: the project (i) evaluated existing routine services delivered to a defined local population; (ii) used aggregate routinely collected referral data rather than identifiable patient-level data; (iii) was designed to improve local pathway performance rather than to generate generalisable new knowledge; (iv) involved no randomisation, allocation, or deviation from standard service delivery; and (v) included no direct contact with patients or service users. These criteria correspond to the HRA definition of service evaluation, which does not require Research Ethics Committee approval or HRA-issued exemption documentation [19].

No formal written exemption document was issued, since none is required in the UK for service-evaluation activity falling outside the definition of research. The classification was made locally by the supervising public-health team and the project was logged within the Directorate’s service-evaluation and quality-improvement records. The work was carried out in line with Blackpool Council’s information-governance policies and with the obligations set out in the General Medical Council’s Good Medical Practice.

All data used were aggregate referral counts held by Horizon, with no patient-identifiable information accessed at any stage. All data were handled in compliance with the UK Data Protection Act 2018, the General Data Protection Regulation, and the NHS Caldicott Principles. No identifiable patient or service-user information is included in this manuscript; the proposed pathway is illustrated only at the system level.

## 3. Results

### 3.1 Epidemiological context

Blackpool carries a disproportionate and persistent burden of drug-related harm. Between 2019 and 2021, 76 deaths from drug misuse were registered, yielding an age-standardised rate of 19.4 per 100,000 population, which is approximately four times the England rate of around 5.1 per 100,000 [1,8]. More recent ONS releases show rates continuing to rise, with Blackpool reporting an age-standardised rate of 30 per 100,000 in 2020–22 and 32.4 per 100,000 in 2022–24, each showing Blackpool as the local authority with the highest drug-related mortality in England [1,2]. Blackpool’s rate of drug-related hospital admissions has similarly remained several multiples of the England average [8].

These outcomes reflect a wider pattern of entrenched deprivation. Eight of the ten most deprived neighbourhoods nationally were located within Blackpool on the 2019 Indices of Deprivation, and the authority was ranked first overall on both the rank-of-average-rank and rank-of-average-score measures [4,5]. Male life expectancy at birth is 73.1 years, the lowest of any English local authority, and there is an 11.2-year intra-authority range between the most and least affluent wards [6,7]. Two of the main contributors to the life-expectancy gap, both at national and at Blackpool level, are smoking and alcohol; drug misuse acts as an accelerant across several of the Core20PLUS5 priority clinical areas, including severe mental illness, cardiovascular disease prevention, and early cancer diagnosis [13].

### 3.2 Existing referral routes

Five established referral routes into Horizon were identified: (i) referral at the point of prison release; (ii) referral by probation or the courts; (iii) referral via the Lived Experience Team (peer-led outreach, originally developed under the Fulfilling Lives programme and subsequently embedded in the ADDER model); (iv) self-referral; and (v) referral from Liaison & Diversion and the ADDER Police team. Stakeholders reported that the prison, probation/court, and Lived Experience routes functioned reasonably well. By contrast, the police/L&D route was described as fragmented: individuals in custody were commonly directed to self-refer after release rather than being transferred directly into assessment, with predictably low uptake and high re-offending among a group known to have exceptional health needs.

### 3.3 Quantitative analysis of police referrals

Across the 47-month evaluation window (December 2019 – October 2023), Horizon received 5,900 treatment referrals in total. Of these, 269 (4.56%) were recorded as originating from the police. The full monthly distribution of police-referral share is summarised in **Table 1**. Police referrals accounted for fewer than 5% of monthly referrals in 30 of 47 months (63.8%); in 17 of these months, the absolute number of police referrals was three or fewer (representing ≤2.5% of the monthly total). Police referrals represented 5–9.9% of total referrals in 16 months (34.0%) and reached or exceeded 10% in only one month of the entire window — December 2022, when 13 of 120 total referrals (10.8%) originated from the police.

**Table 1.** Monthly distribution of police-originated referrals as a share of total Horizon treatment referrals over the 47-month evaluation window (December 2019 – October 2023).

| Police referrals as % of monthly total | Months (n) | % of 47 months |
| --- | --- | --- |
| < 5% | 30 | 63.8% |
| 5 – 9.9% | 16 | 34.0% |
| $\geq 10\%$ | 1 | 2.1% |
| **Total** | **47** | **100.0%** |
| *Aggregate police referrals: 269 of 5,900 total (4.56%)* |  |  |

This pattern is consistent with findings from the national evaluation of Liaison and Diversion by RAND Europe, which demonstrated that although 88% of people seen by L&D services had at least one identified vulnerability and approximately half had drug or alcohol misuse needs, the translation of that contact into sustained specialist treatment engagement was highly variable [14]. It is also consistent with qualitative evidence from the arrest-referral literature, which identifies the credibility of the custody-based intervention, the perceived coercive context, and operational differences between police and treatment providers as recurrent implementation barriers [15].

### 3.4 Policy alignment: Core20PLUS5

Mapping the findings against the NHS Core20PLUS5 framework produced the following alignment. First, neighbourhoods in Blackpool account for a disproportionate share of the Core20 population nationally: eight of the ten most deprived neighbourhoods on the IoD2019, and the authority ranks first among English local authorities on both rank-of-average-rank and rank-of-average-score measures [4,5]. Second, people in contact with the CJS who have problem substance use map directly onto the PLUS inclusion-health groups named in the NHS England specification, alongside people experiencing homelessness and vulnerable migrants [13]. Third, substance misuse contributes materially to four of the five adult clinical priority areas, namely severe mental illness, chronic respiratory disease, early cancer diagnosis, and cardiovascular disease prevention, and exerts a decisive influence on overall (and healthy) life expectancy [13,20].

### 3.5 The proposed unified pathway

Drawing on these findings and on the international evidence base for police-led diversion to treatment, a six-step unified pathway is proposed and summarised in **Table 2**. The pathway retains existing referral routes that function well but replaces the current opt-in, self-report-reliant police/L&D route with an opt-out direct-transfer model supported by multi-agency governance.

**Table 2.** Summary of the proposed six-step unified criminal justice pathway into drug and alcohol treatment in Blackpool.

| Step | Component | Description and key features |
| --- | --- | --- |
| 1 | Initiate Referral (IR) | Opt-out referral from ADDER Police and the Liaison & Diversion team to a single designated Horizon email address. |
| 2 | Initial Assessment (IA) | Triage by the Horizon team within an agreed turnaround time to determine need, urgency, safeguarding, and onward service. |
| 3 | Tailored Treatment Plan (TTP) | Onward referral to the most appropriate combination of services (OASIS, Adelaide Street, Women's Centre, Horizon, Lived Experience Team, Lighthouse, counselling), including medication-assisted treatment where indicated. |
| 4 | Continuous Support (CS) | Connection to community resources for housing, employment, and social needs; case manager assigned where complexity warrants. |
| 5 | Collaboration and Monitoring (CM) | Regular multi-agency review — police, healthcare, social services, housing, community sector — to assess performance, identify improvement, and share learning. |
| 6 | Evaluation and Adjustment (EA) | Routine analysis of referrals, treatment uptake, re-offending, and substance-related prevalence; iterative modification of the pathway. |

#### 3.5.1 Step 1 — Initiate Referral (IR)

Referrals from the ADDER Police team and the Liaison & Diversion team are made to a single designated Horizon email address using an opt-out model: anyone identified in custody as having problem drug or alcohol use is referred by default, with the individual retaining the right to decline engagement. Opt-out designs have been used successfully in other public-health domains (for example, bloodborne-virus testing in emergency departments and prisons) to overcome low uptake driven by stigma, ambivalence, and decisional inertia, and are increasingly recommended for custody-based interventions [21,22].

#### 3.5.2 Step 2 — Initial Assessment (IA)

Referrals are triaged by the Horizon team within an agreed turnaround time. Triage determines treatment need, clinical urgency (including overdose risk and withdrawal), safeguarding issues, and appropriate onward service.

#### 3.5.3 Step 3 — Tailored Treatment Plan (TTP)

The triaged individual is referred to the most appropriate combination of local services, which may include OASIS, the Adelaide Street Service, the Women’s Centre, Horizon’s own key-working and prescribing service, the Lived Experience Team, the Lighthouse Project, and community counselling. Where opioid use disorder is present, medication-assisted treatment (methadone or buprenorphine) is offered in line with NICE guidance and the evidence base demonstrating substantial reductions in overdose mortality and reoffending [23,24].

#### 3.5.4 Step 4 — Continuous Support (CS)

Individuals are connected with community resources addressing housing, employment, and wider social needs, reflecting the whole-systems approach advocated by Project ADDER and the 2021 Drug Strategy [9,11]. A case manager is assigned where complexity warrants.

#### 3.5.5 Step 5 — Collaboration and Monitoring (CM)

A multi-agency steering arrangement, constituted by police, healthcare providers, social services, housing, and community organisations, meets regularly to review pathway performance, identify bottlenecks, and share learning. This mirrors the whole-systems governance structures identified as enablers in the national Project ADDER evaluation [11,16].

#### 3.5.6 Step 6 — Evaluation and Adjustment (EA)

Routine data are collected and analysed on referral volumes, time-to-first-appointment, treatment uptake and retention, re-offending, and (where linkage is feasible) drug-related mortality, with modification of the pathway on a continuous-improvement basis.

## 4. Discussion

This service evaluation provides the first local quantification of the scale of the police-to-treatment referral gap in Blackpool: across nearly four years of routine data covering 5,900 referrals, only 4.56% originated from the police, despite Blackpool having the highest drug-related death rate in England throughout the evaluation window [1,8]. The pattern was remarkably stable: police contributed less than 5% of monthly referrals in almost two-thirds of months and exceeded 10% in only a single month. In a population this exposed to drug-related harm, a referral route that captures only one in twenty contacts represents a significant missed opportunity for secondary prevention.

### 4.1 Interpretation in the light of existing evidence

The findings align with the national evidence base on criminal-justice-referred treatment. The RAND Europe evaluation of the national Liaison and Diversion programme found that, despite near-universal coverage and identification of at least one vulnerability in 88% of those seen (including substance misuse in approximately half), translation into sustained specialist-service engagement was uneven, and the programme’s main measurable benefit was diversion from custody rather than treatment-uptake uplift [14,25]. Earlier evaluations of arrest-referral schemes in London and elsewhere identified similar structural issues: ambiguity about whether the police role was primarily enforcement or harm reduction, the difficulty of establishing service credibility within custody, and the loss of referrals once individuals left the suite [15].

The Project ADDER impact evaluation, published in 2025, reached analogous conclusions at a programme level. It found that ADDER advanced whole-systems working and created new cross-agency relationships, but that implementation was hampered by the COVID-19 pandemic, longstanding budget reductions, and workforce attrition, and that outcomes on drug-related offending and treatment uptake were heterogeneous across the five pilot sites [11,16]. The Blackpool findings reinforce the inference that whole-systems commitment at the strategic level does not automatically produce a well-functioning police referral pipeline at the operational level; the pipeline has to be engineered explicitly.

Internationally, police-led pre-arrest diversion programmes such as Law Enforcement Assisted Diversion (LEAD) in Seattle, the Gloucester Angel Programme, and the Madison Addiction Recovery Initiative (MARI) have demonstrated that it is feasible to embed treatment referral at the point of police contact and that such schemes can increase engagement with community treatment and reduce arrest and overdose rates among participants [17,18,26]. Common features of these models include proactive outreach by treatment-linked staff, a strong opt-out or low-threshold presumption, and explicit, contractually defined referral pathways, all of which are reflected in the proposed Blackpool pathway.

### 4.2 The rationale for an opt-out model

The choice of an opt-out design is deliberate. Opt-in approaches systematically under-recruit populations characterised by stigma, ambivalence, cognitive load at the point of contact, chaotic circumstances, and distrust of authority, which is precisely the profile of people encountered in police custody with problem substance use. Opt-out designs do not remove autonomy: individuals retain the right to decline. They shift the default so that the path of least resistance is engagement rather than disengagement, which is the ethical logic that has underpinned the move to opt-out bloodborne-virus testing in emergency departments and prisons in England [21,22]. Applied to police custody, an opt-out referral to Horizon does not compel treatment; it ensures that treatment is offered reliably and that the individual’s first post-custody contact is a clinical one rather than a void.

### 4.3 Alignment with the Core20PLUS5 framework

The Core20PLUS5 framework is, in this local context, more than a presentational device. It anchors the pathway in a governance structure with defined trajectories, regional oversight, and dedicated 8A-level roles within Integrated Care Systems, providing a route to sustained resourcing and accountability [12,27]. People in contact with the CJS with drug and alcohol dependence fall squarely within the PLUS inclusion-health group, and substance misuse touches four of the five adult clinical priorities. A police-referral pathway that improves access for this group is therefore not a vertical, siloed intervention but a mechanism for delivering against the headline equity commitments of NHS England.

### 4.4 Strengths and limitations

Strengths of the project include the use of four years of complete, routinely collected referral data covering the entire adult population of Blackpool; the explicit alignment of pathway design with both local operational realities and national evidence; and the embedding of the work within pre-existing public-health governance, increasing the likelihood of adoption. Key limitations are that the quantitative analysis is descriptive rather than inferential; that routine data do not capture every police-originated contact (for example, informal signposting that never reached Horizon); that the denominator reflects referrals rather than unique individuals, so some people will be represented more than once; and that the evaluation pre-dates the full post-pandemic normalisation of custody throughput, which may affect generalisability to current operational conditions. The pathway itself has not yet been piloted at the time of writing, so its effect on referral numbers, treatment uptake, re-offending, and drug-related mortality remains to be demonstrated.

### 4.5 Implications for practice and research

For practice, the findings support four concrete actions: (a) implementation of the proposed opt-out referral pathway as the single, default route from ADDER Police and L&D into Horizon; (b) formal designation of the pathway as a Core20PLUS5 local priority within the Lancashire and South Cumbria Integrated Care Board healthcare-inequality plan; (c) embedded data collection capable of measuring referral volumes, time-to-assessment, treatment retention at three and twelve months, and (via data linkage with Lancashire Constabulary) re-offending at twelve months; and (d) investment in the credibility of the in-custody offer, including the visible presence of Horizon and Lived Experience Team members within the custody environment.

For research, a before-and-after evaluation of the pathway with a contemporaneous comparator (for example, a non-ADDER Lancashire custody suite) would provide the most feasible quasi-experimental design. Qualitative work with police officers, L&D practitioners, and service users on the lived experience of the new pathway would complement the quantitative outcomes and help identify the mechanisms of action.

## 5. Conclusions

Despite bearing the highest drug-related death rate in England and exceptionally entrenched deprivation, Blackpool currently derives fewer than one in twenty of its treatment referrals from the police. A unified, opt-out, multi-agency pathway that is anchored in the NHS Core20PLUS5 framework and informed by the evidence from Project ADDER, the national Liaison and Diversion evaluation, and international police-led diversion schemes offers a credible route to narrowing this gap. If implemented and robustly evaluated, such a pathway has the potential to increase treatment uptake, reduce re-offending, and make a measurable contribution to addressing the structural health inequalities that shorten life in this coastal community.

## Acknowledgments

The authors thank the Horizon team, Lancashire Constabulary (ADDER Police), the Blackpool Liaison and Diversion team, the Blackpool Lived Experience Team, and colleagues across the Blackpool Public Health Directorate for their time and expertise during the development of this pathway.

## AI-use declaration

Generative AI tools (Anthropic Claude, Grammarly accessed 2024–2026) were used by the authors on a limited basis to assist with structuring the essay and language editing. No AI tool was used to generate primary data, to perform the data analysis, or to formulate the clinical, policy or pathway-design conclusions of the work. All scientific content, all references, all numerical figures, and all analytical interpretations were generated, reviewed and validated by the human authors, who take full responsibility for the integrity and accuracy of the manuscript.

## Author contributions

Conceptualization, A.O.A., A.G. and J.M.; methodology, A.O.A., A.G., J.M. and A.B.; investigation and data curation, A.B. and A.O.A.; formal analysis, A.O.A. and A.B.; pathway design, A.O.A., A.B., A.G. and J.M.; writing—original draft preparation, A.B. and A.O.A.; writing—review and editing, A.O.A., A.B., A.G., J.M. and T.I.; supervision, A.G. and J.M. All authors have read and agreed to the published version of the manuscript.

## Data availability statement

The aggregate referral data analysed in this study are held by Horizon, the Blackpool specialist drug and alcohol service, and were accessed by the authors under the routine governance arrangements of the Blackpool Public Health Directorate. Aggregate-level summary data are available from the corresponding author upon reasonable request, subject to the Directorate’s information-governance requirements. No patient-identifiable data are available.

## Institutional review board statement

Ethical review and approval were waived for this study because, in accordance with the UK Health Research Authority decision tool, it constituted a public-health service evaluation conducted under the Blackpool Public Health Directorate’s existing clinical-governance arrangements and did not meet the definition of research [19]. The determination of service-evaluation status was made by the supervising Public Health team at Blackpool Council. All data were handled in line with the Data Protection Act 2018, the General Data Protection Regulation, and the NHS Caldicott Principles.

## Informed consent statement

Patient consent was not sought. The project analysed routinely collected aggregate referral data and did not involve direct contact with patients or service users; no patient-identifiable data are reported in this manuscript.

## Sample availability

The author(s) declare that no physical samples were used in this study.

## Supplementary materials

Not applicable.

